# Autistic voice: Sharing autistic children’s experiences and insights

**DOI:** 10.1101/2024.07.22.24310796

**Authors:** Sinéad L. Mullally, Alice E. Wood, Cherice C. Edwards, Sophie E. Connolly, Hannah Constable, Stuart Watson, Jacqui Rodgers, Kieran Rose, Nic King

**Author notes:** The Autistic Advocate, https://theautisticadvocate.com/ (Kieran Rose). **Correspondence:** Sinéad L Mullally.

## Abstract

There is a critical lack of exploration into the first-hand experiences of autistic children in the psychological literature. We sought to address this using baseline data from a wider mixed-methods study. 136 autistic children (mean age=10.35) completed an online questionnaire. Questions explored children’s understanding of autism, their feelings about being autistic in different contexts, and challenges experienced. Quantitative data revealed limited autism knowledge and understanding in some. Challenges included talking about being autistic and self-advocating for needs, especially with non-family members. Children generally recognised both strengths and challenges of being autistic, although concerns about feeling/being different were widespread, and masking common. Strikingly, although most children felt positive about being autistic at home, significantly fewer felt this to be true when around peers or teachers. Using reflexive thematic analysis, four main themes were developed: (1) Overwhelming Experiences, (2) Unsafe People, (3) Sanctuary, (4) Autistic Identity. Overall, the children felt safest at home with family and/or with autistic/neurodivergent/understanding friends, but most unsafe at school with their teachers and neurotypical peers, where victimization was rife. These findings offer valuable insights into the lives of autistic children, and demand we explore how places of education can be transformed into safe spaces for autistic children.

---

> *“You see, you start pretty much from scratch when you work with an autistic child. You have a person in the physical sense – they have hair, a nose and a mouth – but they are not people in the psychological sense. One way to look at the job of helping autistic kids is to see it as a matter of constructing a person. You have the raw materials, but you have to build the person.”* (Chance, 1974).

## Background

Autistic people frequently report feeling misrepresented, stigmatised, dehumanised, and ultimately harmed by traditional autism research (for discussions see Botha & Cage, 2022; Gernsbacher & Yergeau, 2019; Rose, 2020; for wider context see Silberman, 2017), with the above quotation illustrating some of the problematic and harmful perspectives and rhetoric. Whilst the inclusion of autistic people’s first-hand accounts of their own lived experiences could mitigate against this (Botha & Cage, 2022; Saunders, 2018), autistic people have long argued that this does not happen (Holt et al., 2022). This perceived silencing of autistic voices in research has caused enduring mistrust between the autistic and academic communities (Chown et al., 2017; Milton & Bracher, 2013; Milton, 2014; Pellicano et al., 2014), raised ethical and epistemological concerns (Milton & Moon, 2012), and ultimately produced an autism literature that is perceived by autistic people to lack depth and authenticity (Gillespie-Lynch et al., 2017).

Voices of autistic children are particularly absent in the literature. Indeed, a 2016 meta-synthesis found only four peer-reviewed publications centred on first-hand experiences of autistic children (DePape & Lindsay, 2016). Although autism researchers are now increasing efforts to include the voices of autistic youths, their voices are often overshadowed by adults in stakeholder studies; with themes often principally or solely generated from parent, not youth, data (Horgan et al., 2023; Williams et al., 2019), and professional voices often dominating, due to their privileged role offering summary accounts of autistic behaviours (Petty & Clegg, 2025).

Hearing broad and varied lived experiences of autistic children is important to help stakeholders, including researchers, parents, teachers, and clinicians to move from neuronormative narratives, which are discordant with experiences and priorities of autistic people (Leadbitter et al., 2021), and towards an understanding of autism as “a way of being which is complex and dynamic” (Phung et al., 2021, p. 11).

### Rationale for Study

Only a small number of qualitative studies have explored autistic young people’s self-perception and interpretation of their diagnosis. Early findings reported that autistic young people (aged 9-16 years) seek to distance themselves from their diagnosis, avoid finding out more about their diagnosis, demonstrate a preference for non-disclosure of their diagnosis with others for fear of being singled out as ’not normal’, and struggle to identify any benefits to having a diagnosis in the first instance (Ruiz Calzada et al., 2012).

More recent studies exploring the views of autistic adolescents reveal a more nuanced picture, with participating adolescents often appreciating their autistic strengths and perceiving autism as integral to their identity whilst simultaneously describing autism-related challenges and a personal feeling of disconnection from the world (Berkovits et al., 2020; Trew, 2024). Moreover, wider societal stigma surrounding autism, and assumed incompetencies by others, have been described as negatively influencing autistic adolescents’ interpretation of their diagnosis, their self-understanding, and their self-perception (Jones et al., 2015; Mogensen & Mason, 2015; Trew, 2024). It is also clear that autistic young people often assimilate how others perceive their autism into their emergent identities and, as they are keenly aware of teachers’ and school peers’ often negative perception of autism (Billington et al., 2024; Cohen et al., 2022; Horgan et al., 2023; Humphrey & Lewis, 2008; Williams et al., 2019), they form inherently negative autistic identities such as being perceiving oneself as ’retarded’, as ’a freak’, or as having a ’bad brain’ (Humphrey & Lewis, 2008; see also Williams et al., 2019).

Relatively little empirical research has focused on how autistic children perceive, understand, and experience their autism. The limited evidence available suggests that many autistic children possess only a partial understanding of what autism means and often hold negative views about being autistic (Ruiz Calzada et al., 2012). This is concerning, given findings from the adult literature that a positive understanding of autism can help buffer the effects of stigma (Botha et al., 2020), support the development of a positive autistic identity, and serve as a protective factor against mental health difficulties (Cooper et al., 2023; Cooper et al., 2017). Conversely, low personal acceptance of autism has been linked to increased depressive symptoms (Cage et al., 2018).

Not understanding one’s own autism diagnosis, and having insufficient information and knowledge about autism, can make autistic young people reluctant to share their autism diagnosis and their autistic experiences with others, including with their friends (Crompton et al., 2023; Ruiz Calzada et al., 2012). It likely also impedes sense-making of personal lived experiences by denying children the opportunity to better understand themselves and their differences in an informed way (Crane et al., 2021; Oredipe et al., 2023); ultimately resulting in deep suffering (Davidson & Henderson, 2010; Prince-Hughes, 2005).

### Aims of the Study

Against this backdrop, we report baseline data from a larger repeated-measures study that sought to explore potential benefits of autistic-led autism psychoeducation courses for autistic children. We include qualitative and quantitative data from 136 autistic children (aged 8-14 years) prior to commencing the psychoeducation course. A mixed-methods approach was used to explore the children’s understanding of their autism, their feelings about being autistic, talking about being autistic, and self-advocating for their needs, and whether these feelings and experiences differ across different environments (i.e., at home, when out-and-about with friends, and at school). Although primarily designed to assess how these parameters changed following engagement with the course, the study’s design provided access to a large and uniquely uncircumscribed dataset (i.e., the baseline dataset), offering insights into broad, dynamic, diverse, and complex lived experiences of autistic children currently growing up in the UK.

For reasons described above, we anticipated that participating children would have a limited knowledge and understanding of autism and hold either negative or ambivalent views about being autistic. We also expected that many children would report challenges in their peer relationships and may report loneliness, based on previous findings showing that autistic children are often excluded by their peers from an early age (Stagg et al., 2023) and report lower-quality friendships than their non-autistic peers (Bauminger & Kasari, 2000). However, we acknowledge that loneliness and connection must be interpreted with care, as autistic young people may experience belonging in ways not always captured by conventional measures (Lisboa White et al., 2024).

Informed by findings that teachers often perceive autistic students as more problematic and report less warmth and closeness in these relationships (Blacher et al., 2014), we hypothesized that the children would report difficulties in their relationships with teachers, and that these relational challenges with teachers, combined with peer rejection, could shape how children feel about being autistic.

Given consistent findings that autistic students face significant sensory challenges within school settings (Connolly et al., 2023; Jones et al., 2020), we expected that sensory environments would further contribute to school-related stress and wondered how such experiences might influence autistic identity development in that context.

Finally, given the relative absence of research exploring autistic children’s experiences outside of school, particularly within the home, we sought to investigate whether the children’s feelings, self-expression, and language they use when talking about being autistic with others, will vary across different environments.

## METHODOLOGY

### Participants

Participants were recruited as part of a wider study evaluating usefulness of an autistic-led autism psychoeducation course (NeuroBears) that aims to provide post-diagnostic support for autistic children and families. This study had three phases: Baseline phase (where knowledge of autism and feelings about being autistic were assessed prior to engaging with NeuroBears), Course phase (where participants accessed the NeuroBears course), and Post-NeuroBears phase (where baseline questions were repeated, with additional questions specific to NeuroBears). This paper presents data from the children’s Baseline responses only.

In total, 141 autistic children (with safe adults) volunteered to participate. A safe adult was defined as the adult the child felt safe completing NeuroBears with. In all cases this was the child’s parent(s). Data from five children were excluded; four due to completing <50% of the baseline questionnaire; and one for falling outside the remit of the study; leaving a total of 136 participants (for full details see Supplemental Material File 1: Figure S1).

Children were aged between 8-14 years (see Table 1 and Figure 1A). All participants were current residents of the United Kingdom (Figure 1B). Socioeconomic status (SES) of the children was interpreted using the Index of Multiple Deprivation (IMD) (Noble et al., 2019) i.e., area-based deprivation indices based on a combination of seven different types of deprivation (which ranks the population from most to least deprived). The mean IMD decile was less deprived than the UK average (see Table 1 and Figure 1C).

**Figure 1:**
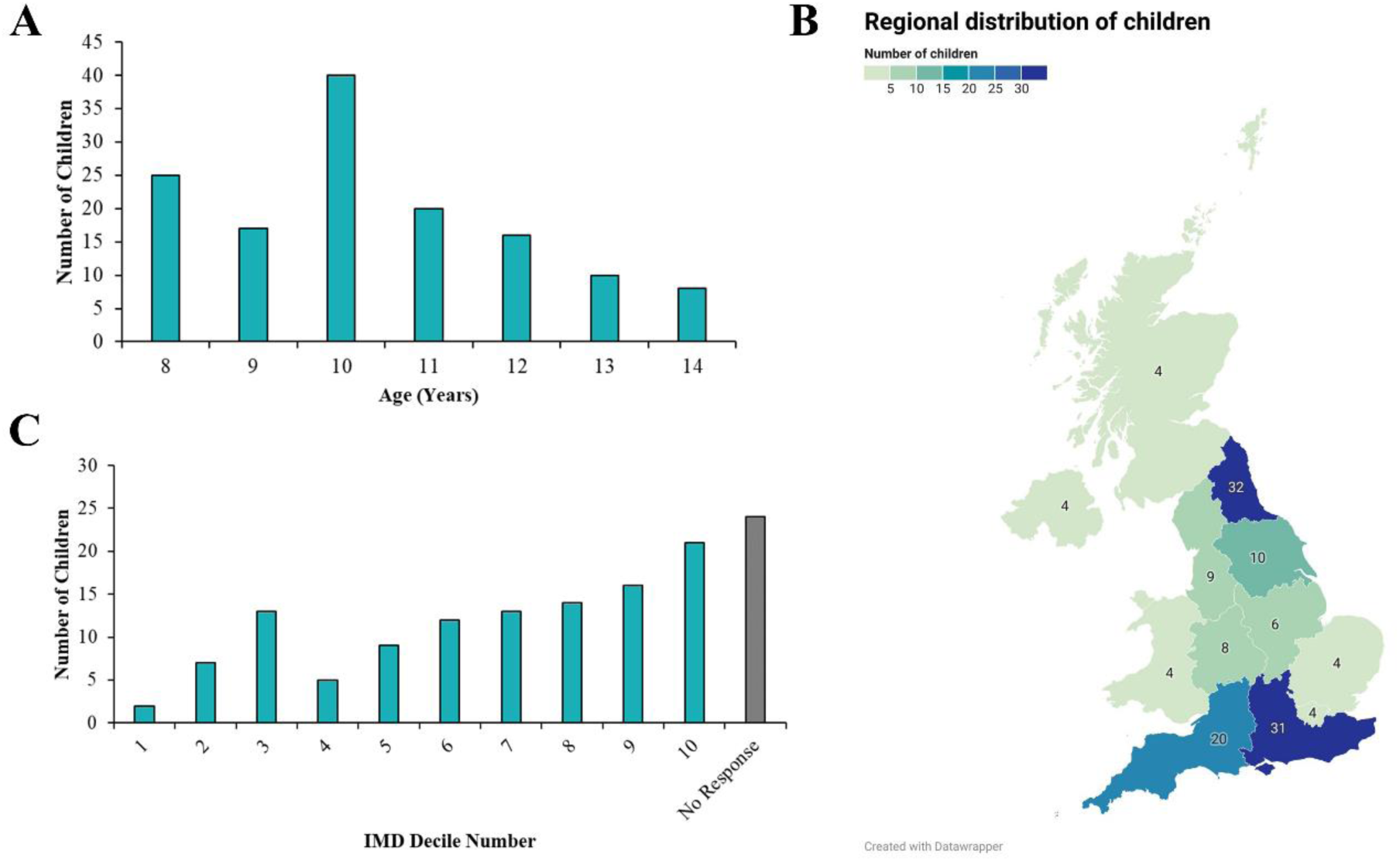
A: Age Distribution of Children (n = 136). B: Regional Distribution of Children (n = 136). C: Index of Multiple Deprivation (IMD) Deciles and Socioeconomic Status (SES) Overview for participants residing in England. Deciles reflect the most deprived neighbourhoods (1) and the least deprived neighbourhoods (10) in England.

**Table 1.** Demographics and Diagnostic Information.

|  |  |
| --- | --- |
| **Age** | Mean = 10.35 years<br>StDev = 1.73 years<br>Range = 8-14 years |
| **Sex Assigned at Birth** | Female: 71 (52.2%)<br>Male: 65 (47.8%) |
| **Gender Identity** | Female: 69 (50.7%)<br>Male: 63 (46.3%)<br>Non-Binary: 2 (1.5%)<br>Self-Described: 1 (0.7%)<br>Prefer Not to Say: 1 (0.7%) |
| **Ethnicity** | White: 128 (94.1%)<br>Mixed/Multi-ethnic: 8 (5.9%) |
| **IMD (Decile)** | Mean = 6.69<br>StDev = 2.69<br>Range = 1-10 |
| **Autism Diagnosis** | 86% formal diagnosis<br>14% self-identification |
| **Time Since Autism Diagnosis** | Mean = 2.06 years<br>StDev = 2.29 years<br>Range = 0-9 years |
| **Autism Status of Reporting Parent** | Autistic: 36% (n=49)<br>Possibly autistic: 37.5% (n=51)<br>Not autistic: 26.5% (n=36) |
| **% of Participants with $\geq 1$ Other Autistic Family Member** | ~91.90% |

As per inclusion criteria, all children were autistic and co-occurring conditions prevalent (see Supplemental Material File 2: Table S1). Given autism’s high heritability (Sandin et al., 2017), we gathered information on whether the children had other autistic family members. 73.5% of parents stated they were either autistic or possibly autistic (see Table 1). Of the 26.5% (n=36) of the parents who are not autistic, 69.4% (n=25) stated it was possible someone in their child’s family was. Hence, it was likely that 91.9% of the cohort had at least one other autistic family member.

### Research Ethics and Language

This study was approved by the Faculty of Medical Sciences Research Ethics Committee, part of Newcastle University’s Research Ethics Committee. Consistent with endorsements from autistic adults in the United Kingdom (Kenny et al., 2016), we use identity-first language within this paper (e.g., ‘autistic child’ rather than ‘child with autism’). However, as the children’s words have been reproduced, some quotes include the use person-first language and/or contain grammatical errors.

### Design

The study used a concurrent embedded mixed-methods, within-participants design, in which both qualitative and quantitative data were collected from both the children and their safe adults, both before and after they engaged with NeuroBears (see Supplemental Material File 1: Figure S1 for further details). As this paper centres on the child’s voice, all data from safe adult questionnaires will be reported elsewhere. Moreover, as this paper focuses solely on children’s lived experiences and opinions <u>prior</u> to commencing the NeuroBears course, only the baseline child data is reported here; alongside demographic and health information (provided separately by consenting parent/guardian).

### Materials

Questions and response options were developed by researchers at Newcastle University (SM, SC, HC, AW, CE, with advice from NK). Questionnaires were distributed via Qualtrics. The child baseline (i.e., pre-NeuroBears) questionnaire focused on children’s understanding and feelings about being autistic, how they communicate with others about their autistic experiences, how they feel when doing so, and how brave they feel to self-advocate (see Supplemental Material File 3). Free text comments informed qualitative analysis. Summary and formal statistics are reported here for key quantitative questions. For details on NeuroBears (www.pandasonline.org) see Supplemental Material File 1: Figure S2.

### Procedure

Participants volunteered by responding to recruitment posts on PANDAS online social media pages and the Newcastle University Cognitive Development Lab’s Facebook page. Participants were recruited via posts on PANDAS social media and the Newcastle University Cognitive Development Lab’s Facebook page. Parents received initial questionnaires by email, which children could complete either independently online or via a video call with researchers. The video call option supported children who might struggle with the literacy demands of the online format or preferred to respond verbally. Independent completion was available for those who might find live interaction with unfamiliar adults challenging. This flexible, choice-based approach was advocated by autistic parent-researchers and aligns with emerging evidence supporting individualised methods to enhance autistic children’s participation in research (McGregor et al., 2025).

Children were also invited to use alternative communication methods, though none chose to do so. In video sessions, the researcher shared their screen and read each question and response option aloud. All children in these sessions chose to respond verbally, with answers transcribed in real time by the primary researcher. A second researcher attended all sessions to ensure accurate documentation of both responses, and any clarifying dialogue initiated by the child. In total, 13 children completed the questionnaire via video call and 123 completed it independently online.

### Community involvement statement

NeuroBears was developed independently by autistic parents of autistic children, in collaboration with autistic young people, to address the lack of non-stigmatising post-diagnostic support for autistic children. Its co-creator (NK) initiated a research collaboration with the lead author (SM), leading to the co-construction of research aims and methods alongside clinical advisors (JW, SW), research assistants (AW, CE, SC, HC), and co-creator KR. Hence, the research team consisted of autistic advocates and educators, alongside autistic, neurodivergent, and neurotypical academics and clinicians. Multiple members of the research team also parent autistic/neurodivergent children and all members of the research team, including the creators of NeuroBears (NK and KR), share authorship on this manuscript.

To centre participant voice in the interpretation and dissemination of findings, a live online “results watch party” was held via Zoom (April 19, 2024). All child participants and their families were invited via email. SM, AW, and CE presented child-friendly findings using slides, with space for questions, discussion, and a movement break. Discussion topics raised by the children and their families included naming NeuroBears characters (children), expanding the animal cast (children), the act of assigning positive or negative valence to certain emotions (family), dissemination plans (both) and future research directions (both). Following on from this session, a draft of the manuscript was emailed to all families who took part in the research. All child feedback was incorporated into dissemination plans and into an updated version of NeuroBears (2.0). Parent feedback on language choices, dissemination places, and future research directions will guide future writings and research plans.

## Data analysis

### Quantitative Data

Bar and radar charts summarise key quantitative child data. Wilcoxon signed rank tests were used to explore the difference between responses to those questions repeated across two contexts. Related-samples Friedman’s analysis of variance by ranks with Dunn-Bonferroni post hoc tests were used where more than two contexts were compared.

### Qualitative Data

We used Braun and Clarke’s (Braun & Clarke, 2006, 2021) style of reflexive thematic analysis (RTA) to analyse children’s free-text responses. RTA was selected to prioritise depth of meaning and attend to participants’ underlying experiences and interpretations. Code and initial theme names were generated at a semantic level meaning that the surface meaning of the what the children told us was taken at face value. A critical realist paradigm was subsequently adopted during the theme development and analysis stages (Fletcher, 2017), providing a framework that acknowledges both the reality of participants’ experiences and the interpretative role of the researcher. Hence, this added explanatory depth by permitting the researchers to make inferences about underlying social structures which may have influenced the children’s shared experiences. By integrating semantic coding with critical realism, we therefore sought to respect the children’s voices whilst also seeking to explore underlying patterns in the data, interpret meaning, uncover mechanisms, and engage with multiple, subjective perspectives. All members of the research team consider autism, and neurodivergence more widely, in a manner consistent with the neurodiversity paradigm in which all neurotypes are equally valued (Chapman, 2019). This, and our collective lived experiences, inevitably influenced our critical realist interpretation and final theme refinement. For further information on the analysis process see Supplemental Material File 4.

### Data availability

Data supporting the study findings is available from the corresponding author upon reasonable request.

## RESULTS

### Quantitative Data

Figures 2, 3, and 4 illustrate distribution of responses to key questions answered by the children (for completeness see also Supplemental Material File 1: Figures S3-S8). As evident from these figures there was considerable distribution of responses across the cohort. However, clear patterns were also evident.

**Figure 2:**
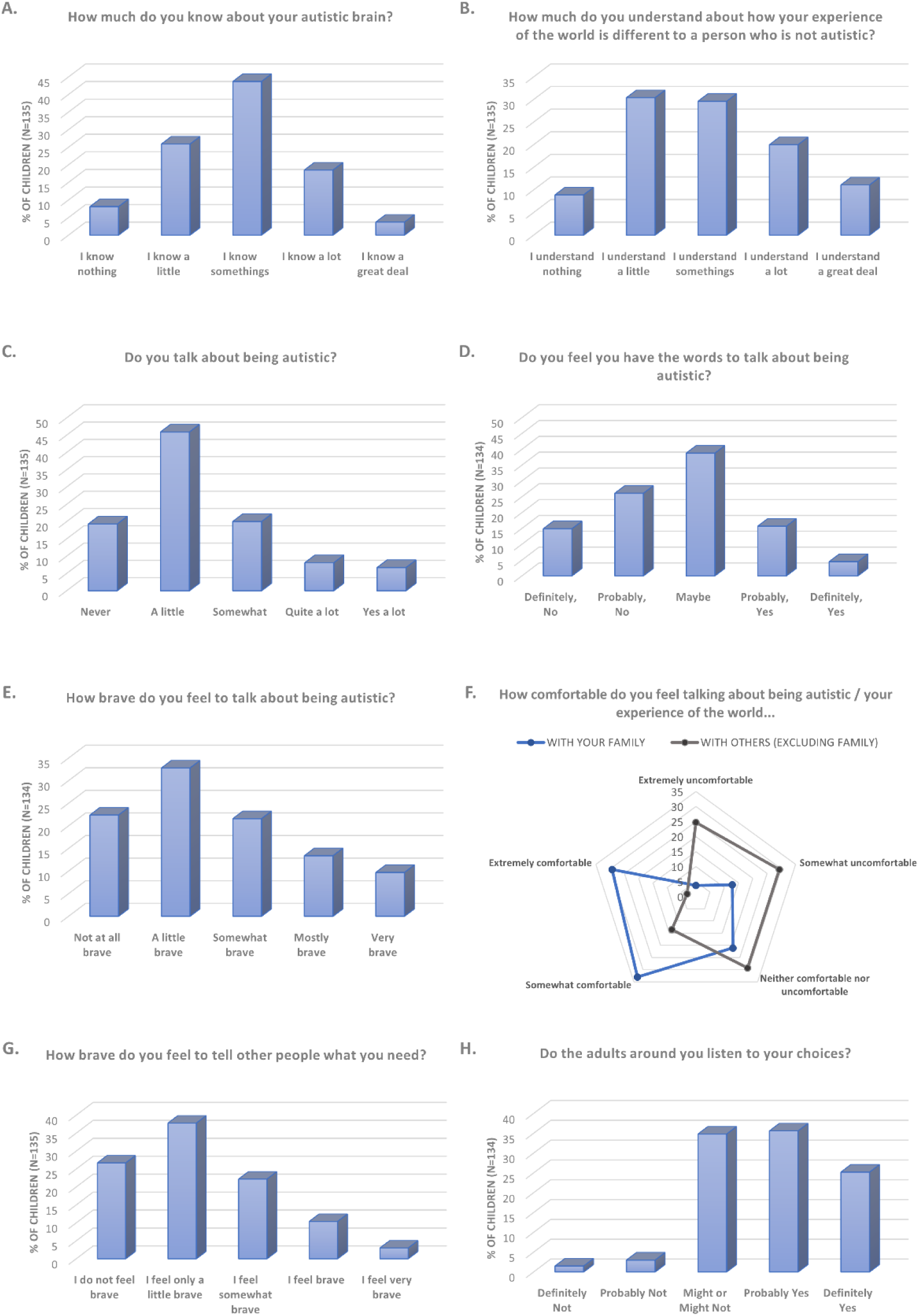
Distribution of the children’s responses to the individual questions (A-H)

**Figure 3:**
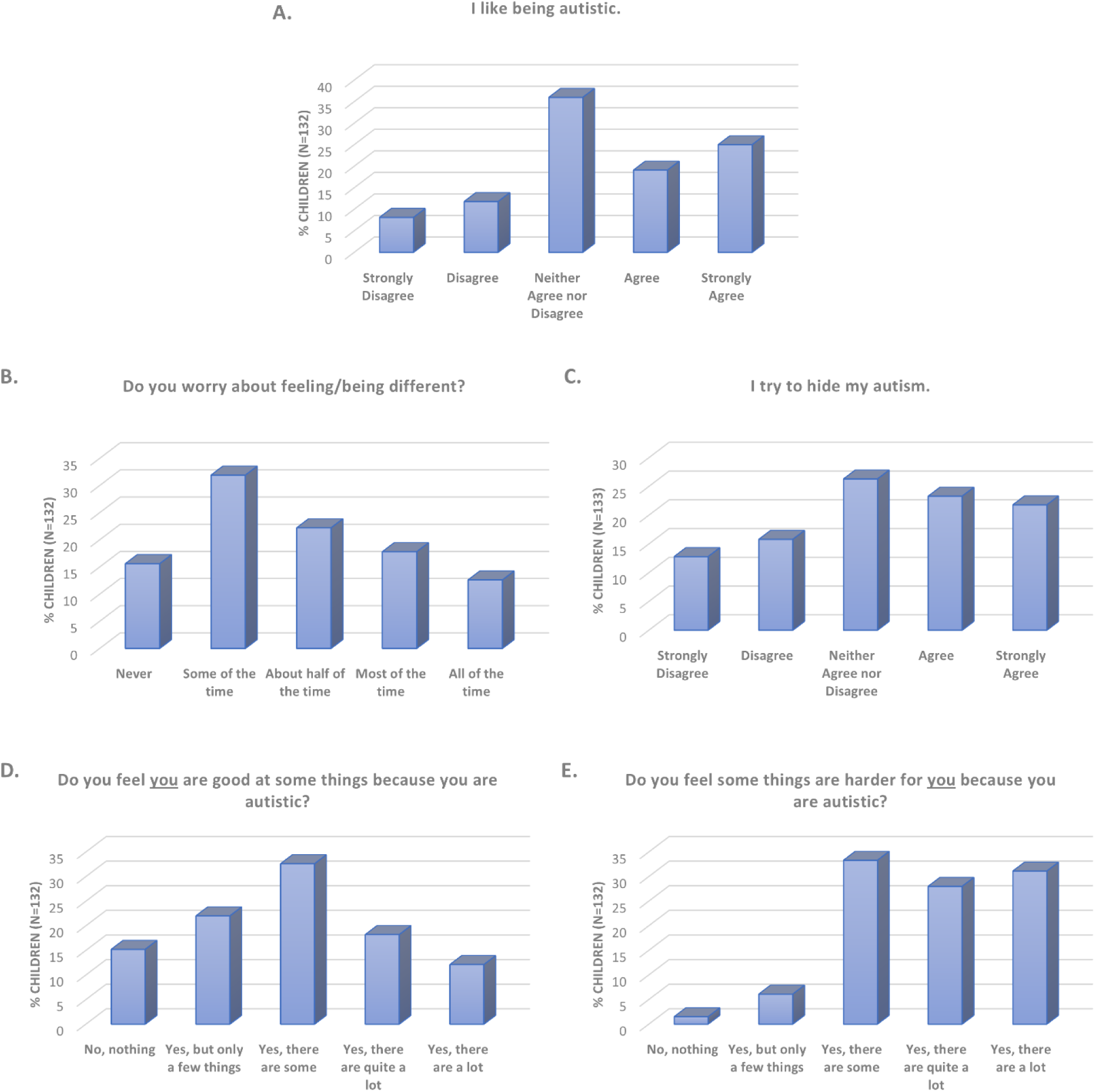
Distribution of the children’s responses to the individual questions (A-E)

**Figure 4:**
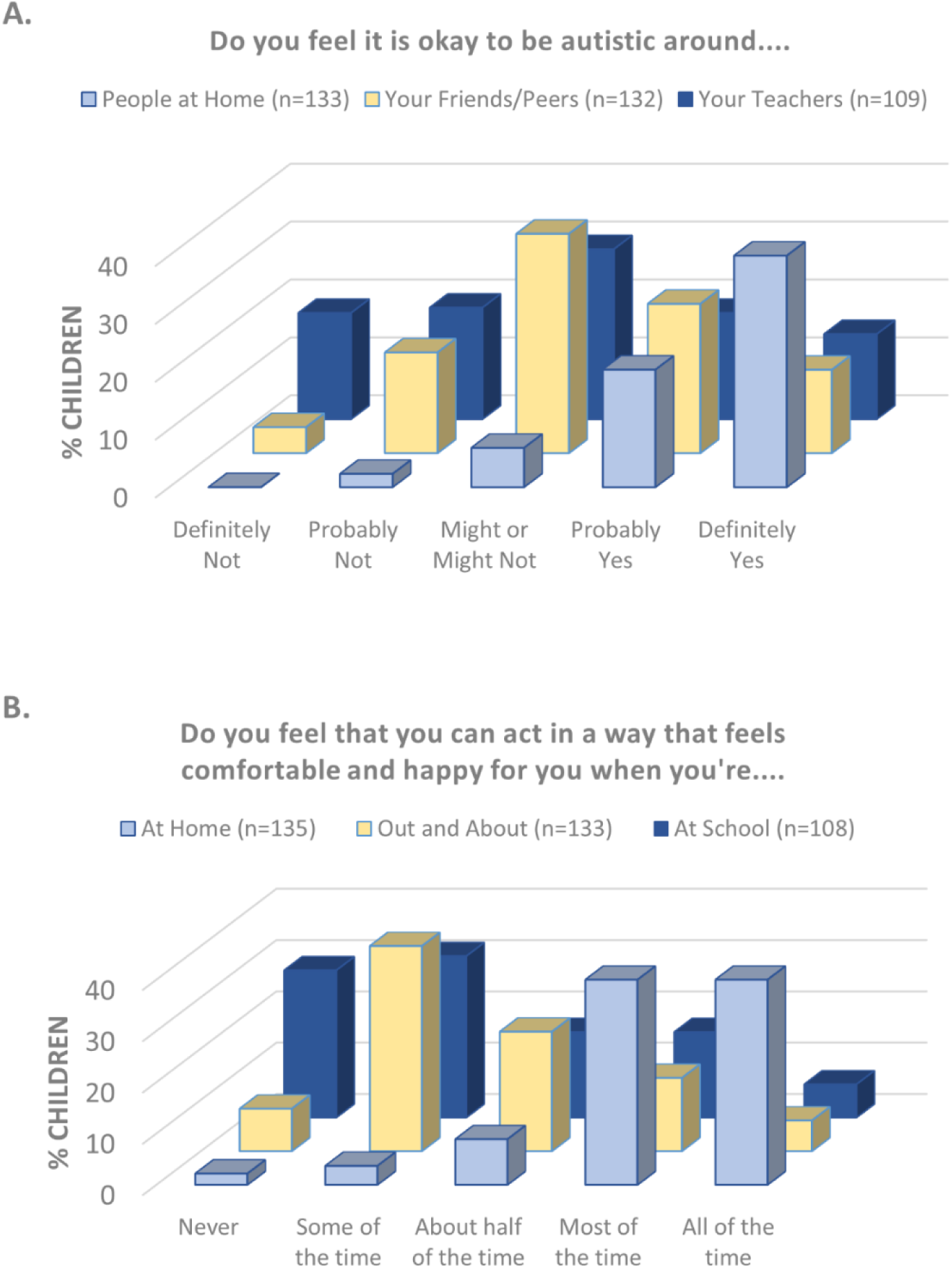
Impact of context on children’s perceptions of (A) whether it is okay for them to be autistic, and (B) whether they feel they can act in a way that feels comfortable and happy for them.

Considering children’s understanding of autism - few children reported knowing ‘a lot’ or ‘a great deal’ about being autistic and over one-third reported knowing ‘nothing’ or ‘a little’ (Figure 2A). Similarly, 39.3% of children reported understanding ‘nothing’ or ‘a little’ about how their experiences of the world differed from those who are not autistic (Figure 2B). Only 14.8% of children reported talking ‘a lot’ or ‘quite a lot’ about being autistic (Figure 2C), and only 4.5% of children reported feeling that they ‘definitely’ have the words to talk about being autistic in the first instance (Figure 2D).

Talking about being autistic with others appeared to be something that the children found particularly difficult - with less than 10% of children reporting feeling ‘very’ brave when talking about being autistic (Figure 2E) and almost one quarter (23.1%) of children stating that they ’never’ feel safe or able to talk to others about their autistic experiences (Supplemental Material File 1: Figure S5).

Moreover, 43% of children selected ‘anxious’ to describe their feelings when talking about being autistic, 31.9% selected ‘nervous’, 19.3% selected ‘scared’, and 14.7% selected ‘panicked’. Comparatively words such as ‘excited’ (2.2%), ‘engaged’ (4.4%) and ‘accepting’ (4.4%), ‘happy’ (5.2%), and ‘calm’ (8.1%) were endorsed less, although 22.2% of children selected the more neutral adjective ‘okay’ and 14.1% endorsed feeling ‘comfortable’.

Importantly, it mattered who these conversations were with. With family, most children (62.4%) reported being ‘somewhat’ or ‘extremely’ comfortable talking about being autistic or their experiences, but this reduced to just 16.5% when these conversations engaged non-family members (Figure 2F). This context difference was significant (Z = -8.311, *p* < .001).

Self-advocating needs was also difficult for the children – with only 13.3% of children feeling either ‘brave’ (10.3%) or ‘very brave’ (3%) to tell other people what they need (Figure 2G). This was despite the majority of children (60.8%) noted that adults around them either ‘probably’ or ‘definitely’ listen to their choices (Figure 2H).

Children’s responses to questions about how they feel about being autistic revealed a complex status quo i.e., although more than double the number of children responded positively to the statement ‘I like being autistic’ relative to those who responded negatively (44.1% relative to 19.9%; Figure 3A), most worried about feeling/being different at least ‘some of the time’ (83.8%; Figure 3B) and almost half reported masking their autism (45.1%; Figure 3C). Similarly, while most children (84.9%) recognised they were personally good at things because they are autistic (Figure 3D), almost all children (98.5%) also stated things are harder for them because they are autistic (Figure 3E).

Children’s responses to the questions of whether they believe it is “okay to be autistic” around people at home, around friends/peers, and around teachers also captured complexity; with 91% of children ‘probably’ or ‘definitely’ agreeing with this statement in the home context, but just 40.2% and 26.8% agreeing when around friends/peers or around teachers respectively. This between-context difference was significant (χ2 (2) = 81.79, *p* < .001), with post hoc tests indicating that the children feel it is significantly more acceptable to be autistic around people at home relative to around friends/peers (*p* < .001) or with their teachers (*p* < .001) (Bonferroni adjustments applied) (Figure 4A). There was no significant difference between ‘friends/peers’ and ‘teachers’.

Significant contextual patterns were also observed when the children rated whether they could act in ways that feel comfortable and happy for them when they’re at home, ‘out and about’, or ‘at school’ (Figure 4B) (χ2 (2) = 85.95, p < .001). Post hoc tests found that the children rated home significantly more favourably than either ‘out and about’ (*p* < .001) or ‘at school’ (*p* < .001), but ‘at school’ and ‘out and about’ did not differ.

### Quantitative Data Summary

Across this large cohort of children, feelings about being autistic and understanding of autism varied. However, several clear patterns emerged. For instance, although more children reported liking being autistic than disliking it and many recognised strengths linked to autism, a majority also worried about being different and almost all children reported that things are harder because of their autism. In addition, the majority reported having limited knowledge about autism, and about how their experiences of the world may differ from a person who is not autistic. Moreover, few children felt they had the language to discuss their autistic experiences with others, and talking about being autistic was generally difficult - with many children expressing feelings of anxiety, nervousness, or fear, and only a minority feeling brave or safe discussing their experiences and to self-advocate. Finally, context was very important, with children feeling significantly more at ease talking about being autistic with family than with non-family members and believing that it is significantly more acceptable to be autistic (or to at in ways that feel comfortable and happy) at home than around their teachers or their peers.

### Qualitative Data

#### Overarching Theme: Growing Up Autistic in a “Neurotypical World”

Throughout the children’s narratives, there was a strong sense of not fitting into the “neurotypical” world, of their challenges being poorly understood, and of finding all of this difficult. In the words of Child 121 “I know people see the world different to me and I find it difficult”. Within this overarching theme, we constructed four main themes. Theme 1 (“Overwhelming Experiences”) and theme 2 (“Unsafe People”) encompassed the challenges that the children described to us, including a loss of felt safety in particular contexts, whilst themes 3 (“Sanctuary”) and 4 (“Autistic Identity”) encompassed what the children told us about their search for safety and their developing autistic identities (see Figure 5 for Thematic Map). It was clear in many children’s responses that the children were aware that the challenges they face in everyday life are not typically experienced by those around them, particularly by their peers. The children were also aware that these challenges were poorly understood by the neurotypical people in their lives.

**Figure 5:**
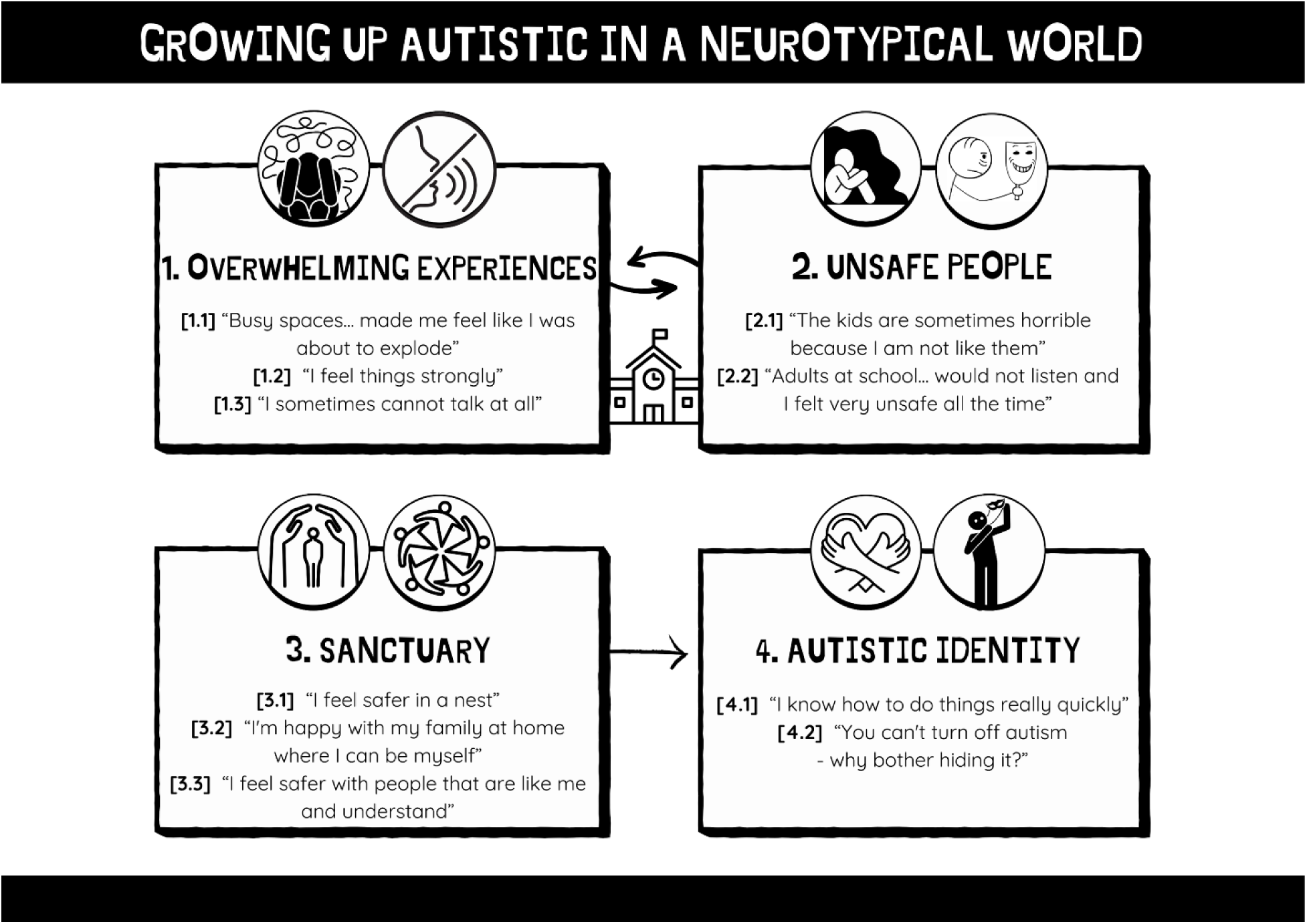
Thematic Map showing the four main themes, alongside the subthemes and overarching theme. The bidirectional arrows between Themes 1 and 2, sitting above an image of a school, have been placed to illustrate how school was experienced by many children as both an unsafe and overwhelming place. The arrow between Themes 3 and 4 highlights the relationship between safety and understanding and the development of a positive autistic identity, as described by some of the children in this cohort.

### Theme 1: Overwhelming Experiences

A prominent theme across children’s accounts was that navigating everyday life often felt overwhelming, frequently leading to dysregulation. This overarching theme comprised three interrelated subthemes: (1) external sensory overwhelm, (2) emotional intensity, and (3) communication shutdown (see Figure 5, Theme 1).

### Subtheme 1.1: “Busy spaces … made me feel like I was about to explode”

Children described frequent sensory overwhelm triggered by everyday environments, especially those outside the home. These included challenges across multiple sensory modalities (e.g., auditory, visual, tactile, and olfactory) with auditory distress most cited. One child shared, “Loud noises scare me” (Child 46), while another explained, “Bright light and noise are way too much. They made me feel hard to focus” (Child 1). Sensory inputs could also cause physical discomfort: “My senses cause me pain sometimes” (Child 57).

Busy or crowded environments were particularly difficult. A vivid example came from one child who recalled, “Busy spaces like the packed boat we were on holiday [made] me feel like I was about to explode” (Child 101). As a result, some children described anxiety about being outside their home environment: “Much harder when not at home, so I worry about going to other places” (Child 32).

### Subtheme 1.2: “I feel things strongly”

Many children described experiencing emotions with marked intensity, which they attributed to their autism. For some, this heightened sensitivity was framed positively: “You’re really kind and caring. You feel things a lot and more emotional” (Child 124). Others expressed that such intensity could contribute to emotional overwhelm and anxiety: “[Being autistic] makes me feel more empathy… [but] makes me more stressed and anxious” (Child 71).

Emotional regulation was often described as challenging, with several children reporting meltdowns when overwhelmed: “Managing my emotions [is harder for me because I am autistic], I have meltdowns when I’m overwhelmed” (Child 40). Some also described difficulties identifying or expressing their own emotions - a characteristic feature of alexithymia: “I’m good at recognising what other people are feeling (but not myself)” (Child 3); making managing intense emotions more challenging and increasing risk of dysregulation.

### Subtheme 1.3: “I sometimes cannot talk at all”

Many children described how overwhelm could inhibit their ability to communicate, especially in unfamiliar or stressful environments. As one child explained, “I sometimes cannot talk at all” (Child 100), while another described: “I get anger inside and my eyes tingle and I can’t tell mummy [what I need] when we’re out” (Child 110).

Some children reported using alternative communication strategies, such as writing: “When I am angry or sad I like to write instead of talk” (Child 8). Others described concealing their distress due to difficulty explaining it, especially in school contexts: “School!!!! School is so hard and changes and when I can’t talk and nobody understands how hard it is for me as I can’t explain it well enough and I just say I’m fine at school but I’m not” (Child 73).

### Theme 2: Unsafe People

A pervasive concern in children’s narratives was the importance of identifying whether others, both peers and adults, were “safe.” Unsafe individuals were typically described as (1) neurotypical peers or (2) adult professionals, particularly in school settings (see Figure 5, Theme 2). Feeling unsafe with peers was most frequently linked to vulnerability to victimisation, whilst feeling unsafe with adults was more typically linked to feeling unheard and misunderstood.

### Subtheme 2.1: “The kids are sometimes horrible because I am not like them”

Children often reported feeling different from their peers due to their autism, which heightened their vulnerability to bullying, exclusion, and social isolation, especially in school. One child explained, “When at school, the kids are sometimes horrible because I am not like them” (Child 93), while another shared, “People think I’m weird as I make faces and noise (stimming), but I can’t help myself” (Child 1). Social isolation was common: “Not really [have] any friends at school (one autistic friend not at school)” (Child 108).

In response, many children described masking their autistic traits to avoid victimisation. For example, “I don’t tell people [I’m autistic] at school because they will bully me” (Child 96), and “[I hide my autism because] some people call me retarded” (Child 11), and “At school I feel like I have to keep them [stims] down as people may see you as something else” (Child 65). Several children noted the emotional toll of masking, with one saying, “I can’t talk at school about it so I just mask and try fitting in then I let it out at home with Mum or sometimes Dad” (Child 73). Others expressed a sense of hiding their authentic selves: “When people make jokes about it, or they don’t know what it’s actually like, I feel like I have to hide it away, and who I actually am” (Child 93).

Feelings of exhaustion from maintaining this façade were frequently reported: “After school… I get very exhausted and I can struggle with this when it gets too much” (Child 3).

Although some children noted that neurotypical peers could be supportive when curious and respectful: “Friends have questions and are interested” (Child 74), many were cautious about disclosing their autism. Trust was built slowly and selectively: “I told two of my friends at school… I don’t want anyone else to know” (Child 3), and particularly difficult within the school context: “[I talk about being autistic] with safe adult [Mum] and friends, everyday. With [other] family, never. Not at school.” (Child 58). Others shared how past betrayals made them wary: “I talked about it with my ex-best friend… he used that to make fun of me. We are no longer friends” (Child 1).

Some children reported difficulty identifying safe peers due to social inference challenges:

“Talking to people and figuring them out are very stressful. I can’t tell whether people genuinely like me or not” (Child 1).

### Subtheme 2.2: “Adults at school… would not listen and I felt very unsafe all the time”

Children frequently described feeling unsafe around adults who did not listen to or understand them, often, school staff. One child reported, “Adults at school when I was there would not listen and I felt very unsafe all the time. This was all adults apart from one” (Child 111). Others were highly discerning about which adults they could trust: “I feel alright talking to SENCO teacher… not around regular teachers” (Child 93), and “I only hang around good adults like my parents, grandparents, and bushcraft mentor” (Child 32).

One child illustrated this difference with a metaphor: “Talking to the right person feels like talking to a fluffy cat or is nice/friendly, make an effort to what I am saying, then I feel that they are the right person. The wrong person feels like a cat that tries to bite you” (Child 69).

Children also reported that their needs were often unnoticed or misunderstood in environments without trusted adults: “If I get overloaded, I don’t think people would notice. People who know me well can pick up on when things are tricky” (Child 79). Some masked in front of adults outside school as well: “I do hide [my autism] if you go to school or with CYPS [Children and Young People’s mental health Services]” (Child 83).

A lack of adult understanding often discouraged children from self-advocating: “I don’t feel brave to tell people at school what I need. I don’t think they listen or get why I’m struggling… [and] I’m scared of being told off” (Child 111). Previous failed attempts reinforced this: “Social worker doesn’t listen [to what I need] even when I was brave enough [to tell them]” (Child 99).

Feeling misunderstood contributed to loneliness and isolation: “If they don’t [understand], it makes me feel alone and sad and isolated” (Child 4). For some, these dynamics made school inaccessible: “At school they don’t listen to things we have asked for… I become tired more easily” (Child 95), and “At school I never felt safe. My mum teaches me at home now” (Child 63).

Despite this, the presence of even one trusted adult could help children manage difficult environments: “I feel safe at home and with close trusted people in unfamiliar or challenging environments” (Child 51). Unfortunately, such trusted adults were rare in many school contexts.

### Theme 3: SANCTUARY

For many children, safety was closely associated with specific people and places that allowed them to regulate, express themselves authentically, and avoid the pressures of social masking (see Figure 5, Theme 3). Home, particularly when cohabited by supportive family members, emerged as a primary sanctuary, alongside other safe spaces and connections with similarly neurodivergent individuals or non-human companions.

### Subtheme 3.1: “I feel safer in a nest”

Home was frequently described as a place of comfort, autonomy, and sensory relief. One child summarised this vividly: “I feel safer in a nest… I can only be happy and feel safe at home” (Child 63). At home, children described greater control over their environment, which allowed them to meet sensory needs more easily: “At home I can use things I need (like chewy necklaces) and move more than at school” (Child 132), and offered reprieve from social expectations: “I worry what people will think about my choices in other places” (Child 107).

Home offered a contrast to school, which was often experienced as overwhelming or invalidating: “I don’t feel comfortable with a lot of the choices I make at school, and at home it is the opposite” (Child 8). Several children referenced the physical comfort of home environments, such as being able to rest, escape from stimulation, or simply be alone when needed: “If things get too much at school, I can’t go somewhere quiet like at home” (Child 36). For some, even small features contributed to sanctuary, such as “Home is better, more comfortable. School chairs are uncomfortable” (Child 132).

Physical containment or enclosed spaces were also comforting for some: “I feel more comfortable in enclosed spaces” (Child 130).

### Subtheme 3.2: “I’m happy with my family at home where I can be myself”

Home was not just a safe space physically but also socially. The presence of safe, understanding people, especially family, was vital. One child stated, “I’m happy with my family at home where I can be myself. I don’t like being out with other people” (Child 63). Many children highlighted that home was where they felt most listened to: “At school, my choices are not listened to. At home, they mostly are” (Child 55).

Being at home also allowed children to unmask and engage in authentic behaviours such as stimming: “I don’t mask at home” (Child 75), and “I can be me at home the most” (Child 111). One child added, “I probably stim more around people I’m comfortable with” (Child 16). The home environment, therefore, enabled emotional regulation and reduced exhaustion.

Within trusted family relationships, children reported feeling more confident to self-advocate: “If I’m with my parents, I would feel comfortable [telling them what I need], but with someone like a normal teacher, I would probably be unable to” (Child 3). They were also more comfortable making choices: “At home I’m very comfortable [to make choices that help me], but at school everything takes a lot of effort” (Child 3). Conversations about being autistic were often limited to the home as well: “Only person I openly talk to about my autistic experiences is my mum” (Child 55).

### Subtheme 3.3: “I feel safer with people that are like me and understand”

Several children identified shared autistic identity, especially with family members, as key to feeling understood. “Different with mum… most likely autistic too, so understand what it’s like when crowds get too busy and it’s too loud” (Child 93). Autistic peers and relatives were seen as easier to relate to and less judgmental: “Less judged by autistic friends” (Child 69), and “I only talk about being autistic to friends I know Will understand (they either are autistic or have a family member who is autistic)” (Child 1).

These relationships fostered mutual understanding and solidarity: “With my friends who have autism or are neurodiverse, it is easier to understand them or the way they are feeling” (Child 93).

Shared humour and collective experiences also contributed to feelings of connection: “Funny jokes with family/friends who are autistic about autism” (Child 93). Some children participated in explicitly autistic spaces, such as art groups or online forums, to access this sense of belonging: “I go to an art class where everyone is autistic. We talk to each other about being autistic. I don’t talk like that in school” (Child 57) and “I also talk about it [being autistic] on TrevorSpace because a lot of autistic people are LGBTQIA+” (Child 16).

For children without such social support, comfort was often found in non-human companions. One child shared: “I don’t have any friends… Toys become my friends and I can play with them” (Child 109). Others described their affinity with animals: “I prefer reptiles. They are like me, they don’t show lots of emotion so I don’t have to read them” (Child 1), and “[Because I’m autistic I’m] good at talking to animals (and understanding what they are saying)”.

### Theme 4: AUTISTIC IDENTITY

Many children reflected on their autistic identity, with responses ranging from acknowledging unique cognitive strengths to articulating challenges in embracing this identity publicly (see Figure 5, Theme 4). The development of a positive autistic identity, closely linked to self-acceptance and reduced masking, was evident in some narratives but often complicated by contextual pressures and a lack of external acceptance.

### Subtheme 4.1: “I know how to do things really quickly”

Children frequently linked their autistic identity to cognitive and sensory strengths. Descriptions included enhanced memory, pattern recognition, and information processing: “Being able to suck in a load of information at one time. Being able to connect that to other pieces of information” (Child 90), and “I know how to do things really quickly” (Child 82). Others highlighted perceptual sensitivity as an asset: “Notice more than others so [I] would be a better detective” (Child 61), and “Not having any sound filters can be helpful” (Child 68). One child humorously expressed heightened awareness as protective: “[You] see things others don’t, [so you are] less likely to get murdered” (Child 30).

Despite acknowledging these strengths, many children also expressed that being autistic often made life more difficult: “[Being autistic is a] harder life for people” (Child 110), and “Everything is harder for me [because I am autistic]” (Child 69). These experiences often overshadowed the benefits, particularly in environments where the children felt misunderstood or unsupported. Thus, while the capacity to name autistic strengths was evident, this did not always translate into the development of a positive identity - particularly where the need to mask to remain safe in everyday life remained strong.

### Subtheme 4.2: “You can’t turn off autism – why bother hiding it?”

Viewing being autistic as being special - with unique skills and characteristics - has been linked with having a positive autistic diagnostic identity in the adult autism literature and with reduced masking (for reviews see Davies et al., 2024; Gray et al., 2024). Here it was evident that a small number of children provided narratives consistent with developing positive autistic identities e.g., “I like being different and I love that I am a bit weird and I love myself” (Child 121) and “You’re unique, you feel like yourself” (Child 126), whilst another child spoke of how acceptance increased their confidence to be openly autistic and resist masking e.g., “You can’t turn off autism - why bother hiding it?” (Child 10).

### Qualitative Data Summary

Four key themes were constructed, all relating to the children’s experiences of growing up as a neurominority. The world beyond the home was often described as overwhelming, unpredictable, and socially unsafe. In these environments, children frequently felt misunderstood or unheard, leading to a pervasive loss of felt safety. To cope, many described the need to hide or mask their autism, which often led to emotional exhaustion, reduced well-being, and anxiety about future interactions.

In contrast, home was typically experienced as a sanctuary, providing sensory comfort, greater autonomy, and emotional safety. Trusted relationships with family members (especially autistic relatives), safe adults, and neurodivergent peers were central to this sense of sanctuary, offering understanding, companionship, and the freedom to unmask and self-advocate.

While many children identified strengths associated with being autistic, such as perceptual sensitivity, deep empathy, or unique ways of thinking, these were often overshadowed by daily challenges and limited opportunities for authentic self-expression. Still, a small number of children described a positive autistic identity, expressing pride in their differences and confidence in being their authentic selves.

## DISCUSSION

This study set out to centre lived experiences of autistic children, addressing a well-documented absence of their voices in traditional autism research (DePape & Lindsay, 2016; Holt et al., 2022). To this aim, we used quantitative and qualitative data provided by 136 autistic children (aged 8-14 years old) at the enrolment point of a longitudinal research project. Baseline questions focused on the children’s understanding of their autism, their varied autistic experiences, their feelings about being autistic (both generally and contextually), and their confidence in self-advocating their needs across contexts.

### The Challenges Growing-Up Autistic a Neurotypical World

A pervasive feature across the children’s narratives was the experience of being overwhelmed by daily life as a neurominority, often due to intense sensory input, complex and uncertain social interactions, and/or heightened emotional sensitivity. As anticipated, this appeared particularly prevalent within the context of the school environment (see Theme 1). Overwhelm often resulted in distress, dysregulation, difficulties describing these experiences to others, and becoming unable to speak. It could also made decision-making difficult, with some children feeling paralysed or unsure of what to do in stressful situations.

Communication difficulties, often intensified by stress and external demands, reflect broader patterns identified in autistic adults, who report similar struggles stemming from both internal factors (like anxiety) and external ones (such as unaccommodating communication partners or overwhelming communicative environments) (Cummins et al., 2020). Difficulties making decisions in challenging environments also aligns with prior research showing that autistic individuals may experience slower decision-making, mental “freezing,” or a tendency to gather excessive information before acting, especially under stress (Luke et al., 2012; Vella et al., 2018). It is also consistent with evidence that autistic adolescents tend to engage in more deliberate and cautious reasoning when making decisions, i.e., they typically take more time to gather evidence and are less likely to jump to conclusions (Brosnan & Ashwin, 2023; Brosnan et al., 2014). Hence, while these traits likely reflect a careful and logical approach, they may also contribute to anxiety and difficulty coping in fast-paced or unpredictable environments. These difficulties may be an overlooked but important feature of being an autistic student for educational professionals to be cognizant of. Moreover, these challenges, alongside the absence of felt safety outside of the family home (see below), ultimately likely contributed to many children describing being autistic as leading to a harder life.

It was also evident from the children’s accounts that they were acutely aware of how they different from their peers, and of the challenges that this presented for them (Theme 2). Autistic children’s awareness of themselves as different to their neurotypical peers is consistent with previous findings (for recent reviews see Horgan et al., 2023; Lynam et al., 2024), as too is the finding that when these differences were perceived as negative, this negativity can become entangled within the children’s developing self-concepts (Humphrey & Lewis, 2008; see also Williams et al., 2019), hence constraining positive autistic identity (Davies et al., 2024). It is for this reason, that many autistic advocates strongly argue in favour of informing autistic children about their diagnoses as soon as possible - as this increases opportunity to increase understand their differences in an informed manner (Oredipe et al., 2023) - hence decreasing the deep suffering described by autistic individuals prior to understanding their diagnosis (Prince-Hughes, 2005).

### Autistic Identity: Emerging Yet Constrained

Developing a positive autistic diagnostic identity has repeatedly been found to benefit psychological wellbeing, and one of the first steps in developing a positive autistic identity is understanding what autism means for you personally (for review see Gray et al., 2024). However, few children in this cohort reported knowing “a lot” about being autistic and over one-third reported knowing ‘nothing’ or ‘a little’. This lack of understanding and knowledge of autism will likely constrain the children’s self-understanding, the development of positive autistic identities, and the ability to describe and advocate for their support needs (Davies et al., 2024; Gray et al., 2024; Oredipe et al., 2023). We also found that only 4.5% felt they “definitely” had the language to talk about being autistic thus, further limiting opportunities to develop their understanding of autism and what it means for them personally through conversations with others.

Despite these challenges, nearly half of the children expressed that they liked being autistic, and a majority recognised personal strengths related to autism, such as heightened perception, creativity, deep empathy, or rapid learning. This did not however always translate into an integrated, confident autistic identity, with positive self-assessments often overshadowed by pervasive concerns about being different from their neurotypical peers (see Theme 2).

Other studies have also found considerable variation between autistic young people in terms of how they view their autism diagnosis - ranging from “oppressive” to “liberating” (Mogensen & Mason, 2015; Trew, 2024), and even a co-existence of conflicting emotions within individual young people (e.g., Jones et al., 2015; Trew, 2024). For instance, Trew (2024) used in-depth narrative interviews with autistic adolescents to reveal that while many embraced their autism as an integral part of their identity, acknowledging strengths such as heightened perception and creativity, they also reported significant feelings of social and emotional disconnection.

Hence, these findings, alongside earlier work, emphasize the nuanced and sometimes conflicting emotions involved in autistic identity development during the childhood and into adolescence. This complex interplay of emotions has also been widely evidenced in autistic adults (for reviews see Davies et al., 2024; Gray et al., 2024). However, to the best of our knowledge, this study is the first to document this in children so young, to quantify the prevalence of these emotions in a large sample of autistic children, and to confirm a preponderance of negative sense-making about being autistic in children.

Moreover, these collective studies suggest that the emergence of autistic pride in autistic children and young people may be constrained - not by a lack of strengths - but by systemic barriers that restrict safe, informed identity exploration and expression. They also underline the importance of context, social acceptance, and the challenges of navigating stigma and masking in shaping how autistic young people understand and express their identities.

### Stigma, Masking, and Burnout

More specifically, the overshadowing of the children’s understanding of positive aspects of being autistic was likely linked to the children’s daily lived experiences i.e., that being ‘different’ makes them targets for bullying, ridicule, and social exclusion. Indeed, only one-third of the children agreed that it is ’probably’ or ’definitely’ okay to be autistic around their friends/peers. These often school-based negative encounters led to social isolation, reduced felt safety, and a need to mask their autistic identity to avoid mistreatment; conditions starkly contrasting with those known to support positive autistic identity development, such as external acceptance and support (Davies et al., 2024).

These findings align with the findings of a longitudinal qualitative study exploring autistic young people’s identity development within mainstream educational settings, which found that many young people experienced tension between a desire to be accepted as “normal” by peers and the reality of feeling different, often resulting in camouflaging or opting out of school-based support to maintain social acceptance (Mesa & Hamilton, 2022).

The desire to hide one’s autism from peers is perhaps unsurprising given previous research showing that eight out of the ten most common stereotypic traits that students associate with autism were derogatory (Wood & Freeth, 2016). The children’s accounts reported here also echo findings from autistic adults, who have reflected that stigma, discrimination, and the framing of autism as a ’bad trait’ often begin in childhood (Botha et al., 2022). Prior research has similarly documented feelings of shame and negativity around autism diagnoses in young people (e.g., Humphrey & Lewis, 2008; Mesa &

Hamilton, 2022; Ruiz Calzada et al., 2012). Consistent with this, many children in our cohort described feeling “anxious”, “scared”, or “panicked” when discussing their autistic experiences.

Furthermore, many children viewed adults (particularly school staff) as emotionally unsafe (Theme 2), with only one-quarter reporting it was ’probably’ or ’definitely’ okay to be autistic around teachers, compared to over 90% for family members. Thus, masking may not only serve to avoid peer victimisation but also to gain acceptance from educators. However, for many, masking carried a significant emotional toll, leading to exhaustion, dysregulation, and a diminished sense of authenticity. These findings align with previous work linking masking with poorer mental health outcomes and burnout (Evans et al., 2024; Miller et al., 2021; Raymaker et al., 2020).

### Teacher bias against autistic students

These findings also raise important questions about teachers’ attitudes towards autism. In a study exploring teachers’ attitudes towards autistic students, participants read two scenarios, one featuring a non-autistic student and a second featuring a seemingly autistic student, and then to indicate their attitudes towards each student (Chung et al., 2015). Results revealed that the participating teachers were more likely to indicate that they would dislike and avoid the autistic student in the scenario relative to the non-autistic student. This aligns with the children’s perception in the present study that being openly autistic is not okay when with their teacher(s).

Further evidence suggests that teachers often report lower-quality relationships with their autistic students, characterised by higher conflict and reduced closeness, when compared with their relationships with their non-autistic students, including their non-autistic students with intellectual disabilities (Blacher et al., 2014; Caplan et al., 2016; Eisenhower et al., 2015; Longobardi et al., 2012).

Higher levels of perceived autistic mannerisms have been shown to be negatively associated with teacher-reported closeness, as measured by statements such as “I share an affectionate, warm relationship with this child” (Blacher et al., 2014). Hence, it is likely astute of the children sampled here to have deduced that it is not conducive to positive relationships with their teachers to be openly autistic in the classroom.

A notable gap in this literature is the lack of research capturing autistic students’ own perspectives. One exception is a more recent study by Blacher and colleagues, which surveyed young autistic children (mean age = 6.6 years; n = 136) in the U.S. (Losh et al., 2022). While 75% believed their teacher liked them, a significant proportion also ‘sometimes’ or definitively felt unheard (36.1%), unfairly treated (40.4%), or frequently in trouble (44.1%) (personal communication with the authors). Although the sample was younger and from a different national context, these findings dovetail with those in the current study and raise concerns.

Future research should directly explore how autistic and non-autistic students perceive their relationships with teachers, particularly whether autistic students feel uniquely compelled to mask core aspects of themselves to gain acceptance from their teacher(s). This is crucial given consistent evidence linking poor-quality student-teacher relationships to worse school adjustment and outcomes (e.g., Baker et al., 2008; Fowler et al., 2008), and, as noted, the association between masking and poorer long-term mental health (Evans et al., 2024; Miller et al., 2021; Raymaker et al., 2020).

### Adults as Unsafe or Emotionally Unattuned

A recurring concern among the children was that adults, often teachers and other professionals, failed to listen, respond empathetically, or understand their needs. This lack of attunement frequently left children feeling isolated, fearful, and unable to self-advocate, often due to fears of being dismissed or punished. Even when they did attempt to express their needs, many felt ignored or misunderstood, further eroding their trust in adults.

### Unsafe and Inaccessible Places

This mistrust extended beyond individuals and into physical environments like school, which many children saw as both emotionally and sensorily unsafe. For many children, unsafe relationships with school staff, peer victimisation (Theme 2), and the sensory and emotional overwhelm often experienced at school (Theme 1), compounded the sense that places of education were unsafe or “a bad space with people that are being sucky” (Child 4); places where attendance caused distress, dysregulation, and exhaustion. As a result, a number of children in the cohort reported being unable to continue in school-based education, with many families subsequently turning to alternatives such as home education when school environments were perceived as persistently unsafe or invalidating. These findings mirror those reported in larger UK-based cohorts (e.g., Connolly et al., 2023; Mullally & Connolly, 2025).

Whilst some children were able to identify some school staff, such as Special Education Needs support staff or specialist teachers, as “safe,” such individuals were rare in school contexts, although when present in a child’s life, could make an otherwise inaccessible space, accessible. Due to the rarity of these relationships, many children felt compelled to mask their autism even around professionals, including teachers and mental health staff, thereby limiting their access to education, support, and opportunities for positive autistic identity development. For similar reasons, unfamiliar places were also described as feared and anxiety provoking.

### The Role of Context and Neurotype in Shaping Safety and Identity

A core contribution of this study lies in its demonstration of the significant role context plays in shaping autistic children’s experiences. Children consistently reported feeling more comfortable discussing their autistic experiences and behaving authentically at home than in school or public settings. Home was frequently described as a sanctuary (Theme 3) from an overwhelming (Theme 1) and unsafe (Theme 2) external world; likely also providing respite from public and anticipated stigma (Han et al., 2022). Statistically significant differences were observed in perceived safety and acceptance across settings, with home rated as the most supportive environment. Home as a place of safety which can provide sanctuary away hostile environments (such as the school environment) has also been briefly described elsewhere (e.g., Fielding et al., 2025; Mesa & Hamilton, 2022), but not, to the best of our knowledge, statistically compared.

Children described home as offering both physical and sensory comfort, a space where their needs were more easily met and where tools for self-regulation were readily available. This aligns with previous findings that caregivers report routines are more manageable at home due to more predictable sensory input and better access to supportive resources (Schaaf et al., 2011). Home environments also typically involve fewer social demands and provide greater autonomy and control (Heyworth et al., 2021).

Central to the sense of safety at home were the relationships children described with trusted family members, particularly those who were also autistic. Autistic relatives were often experienced as allies who intuitively understood the child’s needs and emotional states, serving as safe relational anchors. Neurotypical adults and peers who demonstrated a willingness to understand and reliability were also viewed as safe. Importantly, nearly three-quarters of parents in this study identified as, or possibly as, autistic (Table 1), and children noted how this shared neurotype enhanced their caregivers’ capacity to predict and respond to distressing situations, thereby increasing their felt safety.

Similarly, safety was also described in relationships with autistic or neurodivergent peers, particularly outside of school. These relationships were characterised by shared understanding, freedom of expression, and absence of judgment - contrasting sharply with the children’s descriptions of interactions with school peers and professionals. These experiences mirror concepts from the adult autistic literature, such as autistic social connectedness (deep connections with other autistic individuals) and autistic belongingness (the sense of similarity and resonance among autistic people) (Botha et al., 2022).

Likely relevant is the double empathy problem that recognises that communication between autistic individuals is highly effective and does not suffer the communication issues evident when autistic and non-autistic people communicate (Crompton et al., 2020; Foster et al., 2025; Milton, 2012). Being with autistic family members and friends likely facilitates affective communication, leading to relationships that feel harmonious and safer.

In line with research showing that autistic self-acceptance emerges in environments where individuals feel externally accepted and supported (Davies et al., 2024), schools may benefit from actively fostering these conditions. Drawing on insights from the double empathy literature, schools could work to build affirming environments by supporting autistic peer communities. Initial trials of autistic peer support groups in UK schools have shown promise, with autistic students expressing enthusiasm for such groups, provided they are flexible, inclusive, strengths-based, and affirm neurodiversity (Crompton et al., 2024; Crompton et al., 2023).

### Limitations and strengths

Potentially limiting this research is the format of long questionnaires, that may lead to boredom and shortened answers and selection bias in who could participate in this research. Lack of ethnic diversity within this study is critical to address; as is the necessity to acknowledge the nuanced intersectionality of race, ethnicity, sexuality, gender, socioeconomic factors, and age as factors that create distinct barriers to self-discovery, community-building, and advocacy within the context of autism. The dominant narrative surrounding autism has historically been white and Western-centric (Silberman, 2017), thereby alienating non-white individuals from safe spaces for communal discussion and connection surrounding their multiplied marginalised identities.

Underlying issues of privilege are paramount in understanding this narrative. Individuals with more privileged identities may have the luxury of exploring and acting upon concepts such as neurodivergent authenticity or ’unmasking’, without fear of repercussions. However, members of minority groups may not have the same level of safety and protection when advocating needs or expressing boundaries due to systemic biases and discrimination. It is likely relevant here this cohort had over-representation of families living in least deprived areas of the country. Our participants also all had parents actively seeking a better understanding of autism for them via the act of engaging in this research in the first instance.

## Conclusion

The data presented here highlights the urgent need to change how society, and particularly educational systems, engage with autistic children. The children’s accounts offer a compelling challenge to deficit-based narratives: they do not lack personhood (Chance, 1974), self-awareness and understanding (e.g., Williams, 2010), nor the capacity to produce reliable knowledge on autism (Frith & Happé, 1999). Across contexts, children demonstrated acute social perception, emotional insight, and agency in how they made sense of being autistic, despite frequently encountering stigma, exclusion, and emotional harm.

What emerged most strongly was the central role of safety and acceptance in enabling autistic identity development and self-expression. In affirming contexts, most notably at home and with trusted, often neurodivergent others, children were better able to explore what being autistic meant for them, to feel safe being themselves, to utilise self-regulatory strategies as required, to self-advocate for their needs to be met, and critically, to feel safe being themselves. In contrast, school environments were frequently described as unsafe, overwhelming, and invalidating, contributing to anxiety, dysregulation, communication shutdowns, and widespread masking and its emotional toll. These dynamics reflect not individual deficits but systemic failures.

Moving forward demands more than token inclusion. It requires structural and cultural transformation - rooted in respect, neurodiversity-affirming practice, and the co-creation of safe spaces where autistic children are empowered to express themselves without fear of judgement or harm. This includes expanding access to trusted autistic adults, creating opportunities for autistic peer connection, and rethinking educational norms to better accommodate diverse ways of being.

By centering autistic children’s lived experiences, this study underscores the urgency of dismantling neuronormative systems and narratives, and replacing them with those that affirm autistic identity, autonomy, and personhood.

## Supporting information

see Supplemental Material File 1: Figure S1

(see Supplemental Material File 2: Table S1

see Supplemental Material File 3

see Supplemental Material File 4

## Data Availability

All data produced in the present study are available upon reasonable request to the authors.

