## Supplementary material for "Autistic voice: Sharing autistic children’s experiences and insights": see Supplemental Material File 1: Figure S1

**File 1**

**Figure S1:** Flow-chart detailing the phases of the wider study. The blue box highlights where the data analysed in this paper was acquired.

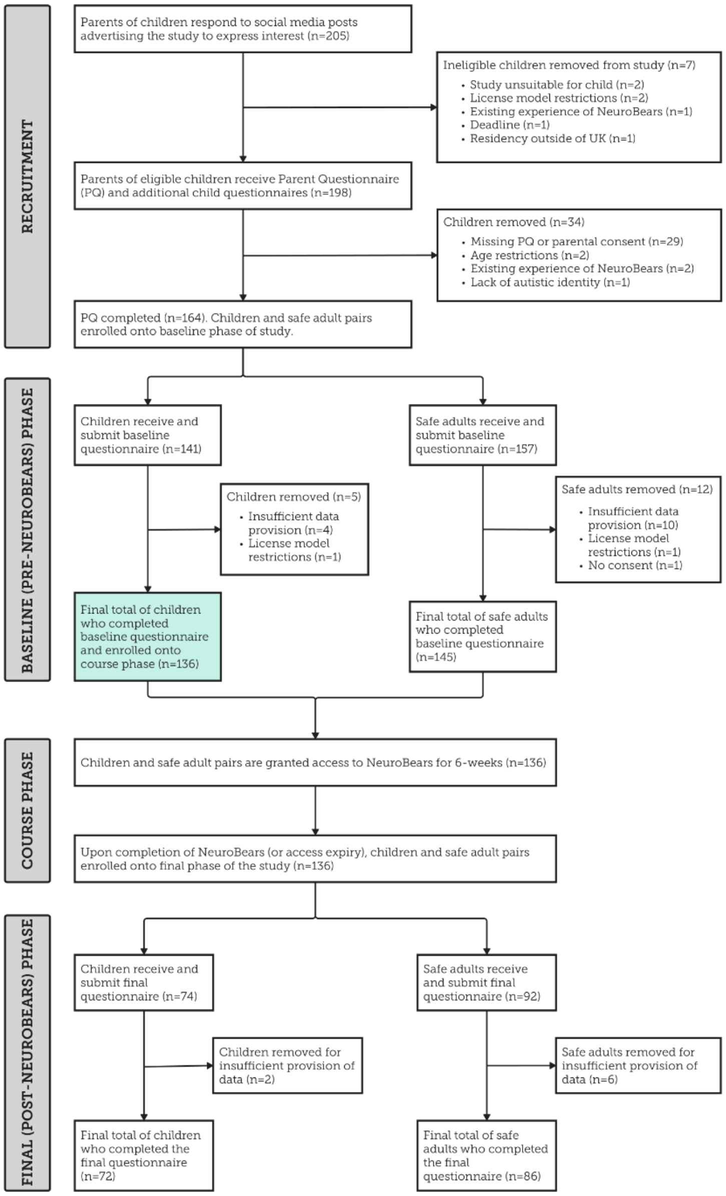

**Figure S2:**

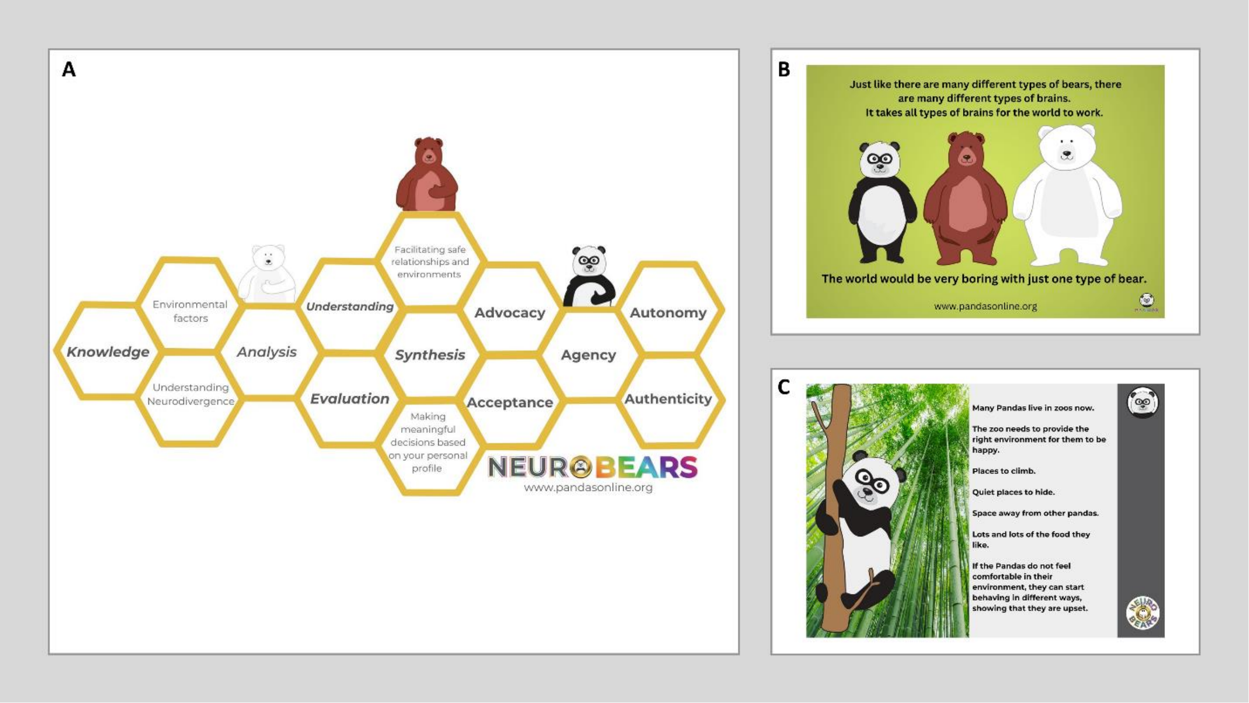


NeuroBears (https://www.pandasonline.org/catalog) is a pre-recorded psychoeducation course all about autism. It was developed by autistic individuals, for, and in collaboration with autistic children and is not affiliated with any health or academic organisation. NeuroBears is primarily aimed at autistic children aged 8-14, who are new to understanding their autism. It aims to educate and increase autistic children's understanding of their autistic experiences, to provide them with context, knowledge, and language (see Panels A-B). In doing so, it seeks to empower autistic children to build the confidence to share these experiences, and to promote the development of a positive autistic identity, and awareness of the wider autistic community to which they belong (see Panel C). It also aims to support the children's safe adult(s) by developing their knowledge of autism, common autistic experiences and co-occurring conditions. NeuroBears contains 12 videos which are under 90 minutes in total (maximum individual video length <12 minutes) and is accompanied by 5 guides/young person and adult workbooks.

**Figure S3: Additional Question in the Pre-NeuroBears Questionnaire**

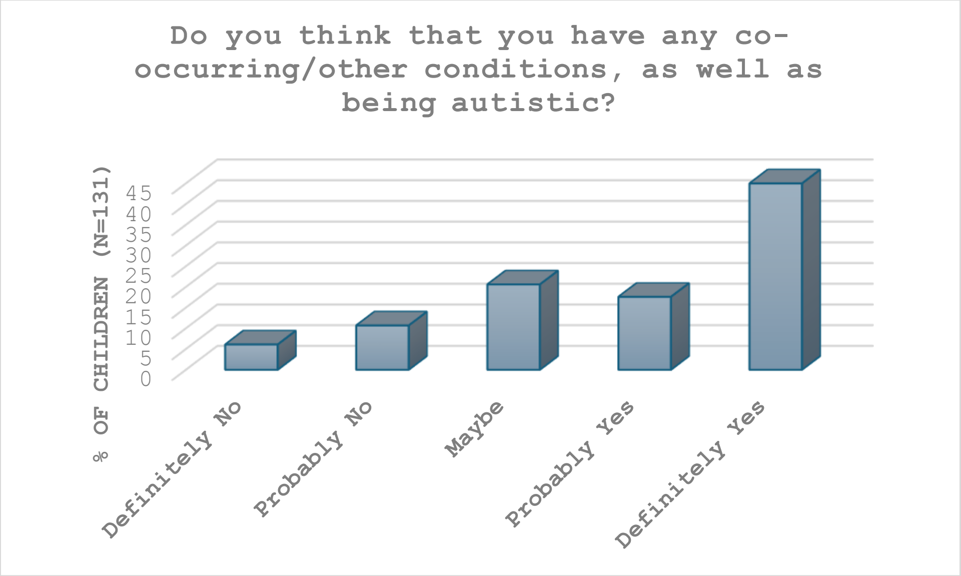


**Figure S4: Additional Question in the Pre-NeuroBears Questionnaire**

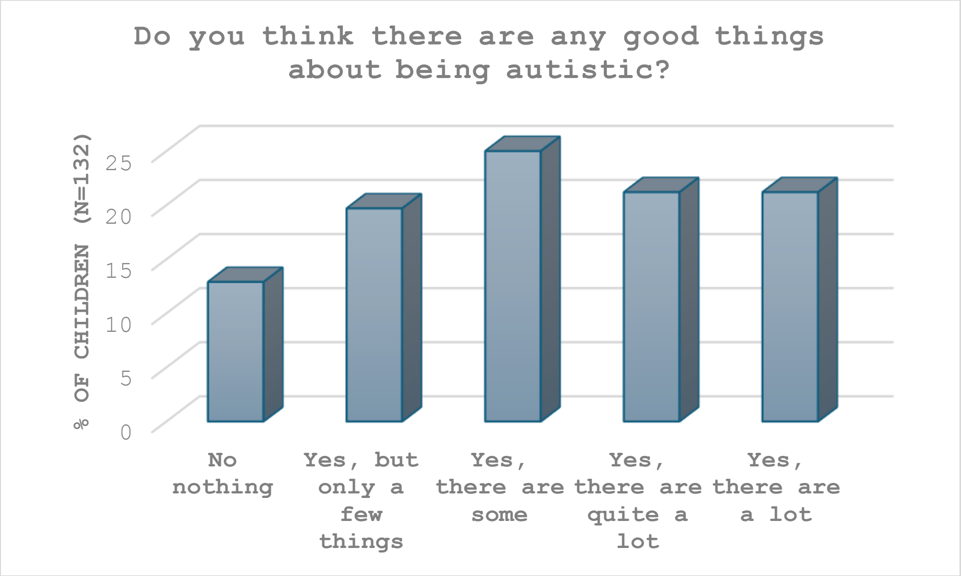

**Figure S5: Additional Question in the Pre-NeuroBears Questionnaire**

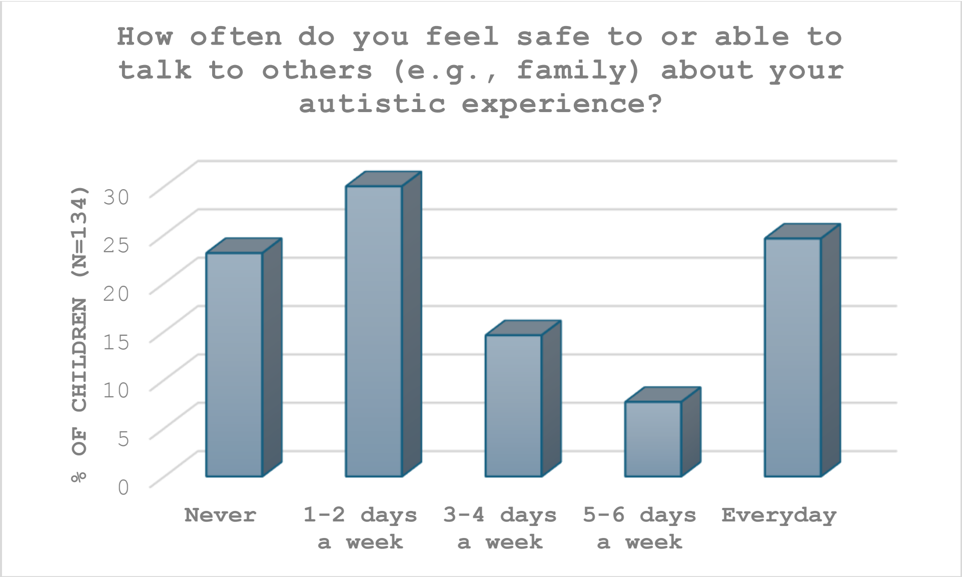


**Figure S6: Additional Question in the Pre-NeuroBears Questionnaire**

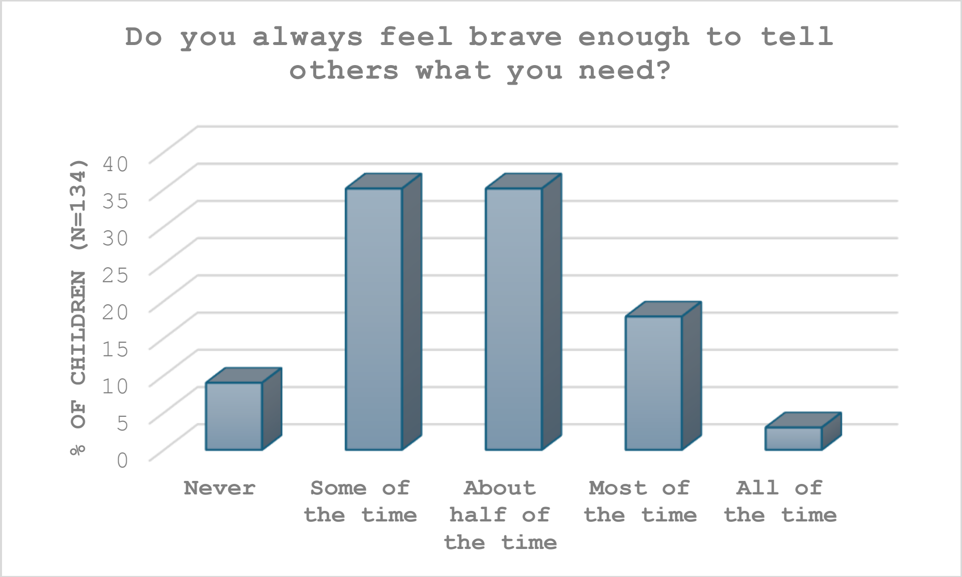

**Figure S7: Additional Question in the Pre-NeuroBears Questionnaire**

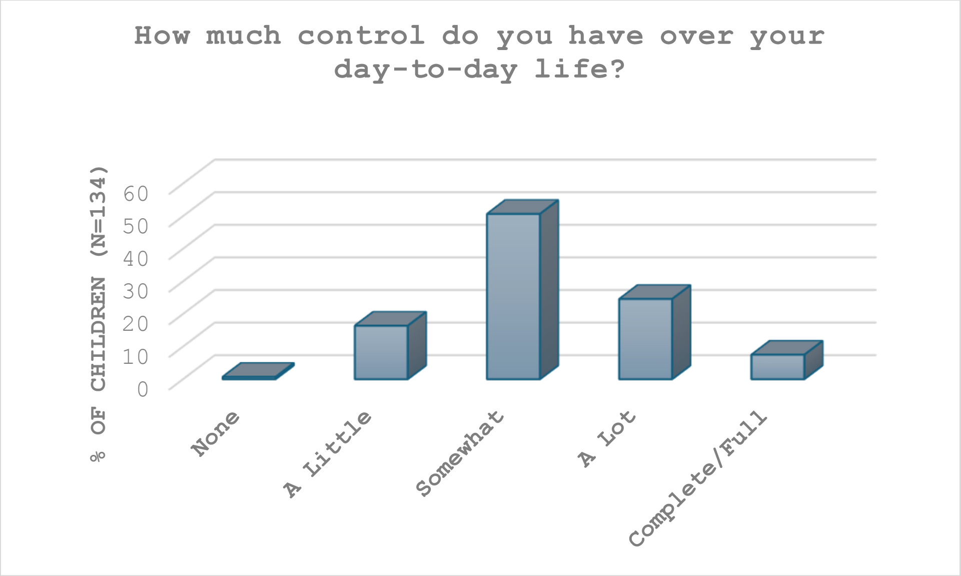


**Figure 8: Additional Question in the Pre-NeuroBears Questionnaire**

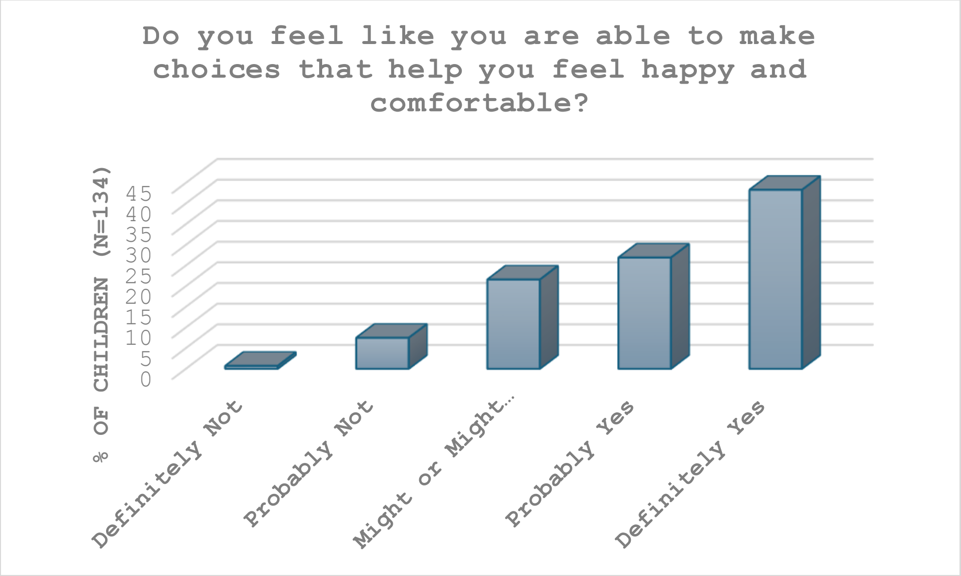
