## Supplementary material for "Autistic voice: Sharing autistic children’s experiences and insights": (see Supplemental Material File 2: Table S1

**File 2**

**Table S1: Summary of neurodivergences within the cohort.**

| Diagnosis | Total Number of Participants (N=136) | Percentage (%) |
|---|---|---|
| **Autism** | 136 (86% diagnosed, 7.4% self-identified) | 100% |
| **Attention-Deficit/Hyperactivity Disorder (ADHD)** | 28.7% diagnosed, 14% on diagnostic pathway, 16.9% self-identified | 61.10% |
| **Sensory Processing Difficulties** | 22.8% diagnosed, 4.4% on diagnostic pathway, 28.7% self-identified | 57.40% |
| **Anxiety** | 19.9% diagnosed, 4.4% on diagnostic pathway, 25.7% self-identified | 52.20% |
| **Emotion Regulation Difficulties** | 8.1% diagnosed, 0.7% on diagnostic pathway, 33.8% self-identified | 44.10% |
| **Pathological Demand Avoidance (PDA)** | 12.5% diagnosed, 5.1% on diagnostic pathway, 19.9% self-identified | 37.50% |
| **Hypermobility** | 14% diagnosed, 2.2% on diagnostic pathway, 14% self-identified | 30.90% |
| **Dyspraxia/Motor Skills Disorder** | 8.8% diagnosed, 1.5% on diagnostic pathway, 11% self-identified | 21.30% |
| **Dyslexia** | 6.6% diagnosed, 2.9% on diagnostic pathway, 9.6% self-identified | 20.60% |
| **Selective Mutism** | 6.6% diagnosed, 1.5% on diagnostic pathway, 10.3% self-identified | 19.10% |
| **Auditory Processing Disorder (APD)** | 2.2% diagnosed, 0.7% on diagnostic pathway, 11% self-identified | 15.40% |
| **Gifted** | 0.7% diagnosed, 13.2% self-identified | 13.90% |
| **Depression** | 1.5% diagnosed, 2.2% on diagnostic pathway, 3.7% self-identified | 8.10% |
| **Dyscalculia** | 2.2% diagnosed, 5.9% self-identified | 8.10% |
| **Speech Difficulties** | 4.4% diagnosed, 0.7% on diagnostic pathway, 2.9% self-identified | 8.00% |
| **Visual Processing Difficulties** | 2.9% diagnosed, 0.7% on diagnostic pathway, 4.4% self-identified | 8.00% |
| **Dysgraphia** | 2.2% diagnosed, 0.7% on diagnostic pathway, 4.4% self-identified | 7.30% |
| **Obsessive Compulsive Disorder (OCD)** | 3.7% diagnosed, 0.7% on diagnostic pathway, 2.9% self-identified | 7.30% |
| **Tic Disorder** | 0.7% diagnosed, 1.5% on diagnostic pathway, 2.2% self-identified | 5.10% |

| | | |
|---|---|---|
| **Other Physical Health Condition(s)** | 2.9% diagnosed, 1.5% self-identified | 4.40% |
| **Language Disorder** | 2.9% diagnosed, 0.7% on diagnostic pathway, 0.7% self-identified | 4.30% |
| **Other Mental Health Condition(s)** | 2.9% diagnosed, 0.7% on diagnostic pathway, 0.7% self-identified | 4.30% |
| **Other Neurodivergence(s)/Health Condition(s)** | 1.5% diagnosed, 0.7% self-identified | 2.20% |
| **Colour Vision Deficiency** | 0.7% diagnosed, 0.7% self-identified | 1.40% |
| **Intellectual Disability** | 0.7% self-identified | 1.40% |
| **Unspecified Learning Disorder** | 0.7% self-identified | 1.40% |
| **Other(s)** | 0.7% self-identified | 0.70% |
