## Supplementary material for "Autistic voice: Sharing autistic children’s experiences and insights": see Supplemental Material File 3

**File 3**

### Examples of the different formats of the free-text questions

#### Example Free-Text Box:

**If you have any other comments on your experiences of talking about being autistic, you can add them here.**

**Also, if your preferred way of communicating about being autistic isn't with speech, you could add this here and tell us more about this.**

#### Example Likert Scale Response Options:

**Using the scale below, please indicate how much you agree or disagree with the two following statements:**

**(Note: You can add any comments about these questions in the boxes below.)**

| | Strongly Agree | Agree | Neither agree nor disagree | Disagree | Strongly disagree |
|---|---|---|---|---|---|
| - I like being autistic. | ○ | ○ | ○ | ○ | ○ |
| - I try to hide my autism. | ○ | ○ | ○ | ○ | ○ |
