## Supplementary material for "Autistic voice: Sharing autistic children’s experiences and insights": see Supplemental Material File 4

**File 4**

**Additional Information: Thematic Analysis Procedure**

AW and CE began data analysis with familiarisation i.e., reading and notating the data multiple times. AW and CE independently coded data, joining back up to review codes and ensure that the coding system was systematically coding all interesting data features. Next, similar codes were grouped into potential themes, which were reviewed and refined by the team in an iterative, collaborative process, until a finalized set of named themes were decided, and corresponding thematic map generated. AW and CE revisited raw data and codes throughout the process to ensure generated themes accurately reflected the data. Following initial review, and acting on reviewer advice, SM (with discussions with the research team) iterated the above process, generating revised themes, theme/subtheme names, and corresponding thematic maps. This latter step was redone following an additional review process.
